# Life Course Sleep Duration Trajectories and Risk and Age at Onset of Parkinson’s Disease

**DOI:** 10.1101/2025.02.25.25322885

**Authors:** Yi Fang, Rebecca Hardy, Kristine Yaffe, Simon J. Little, Caroline M. Tanner, Yue Leng

## Abstract

**Importance:** Investigating associations between life course sleep duration and Parkinson’s disease (PD) may help clarify the role of sleep in early detection and/or prevention of PD.

**Objective:** To characterize life course sleep duration trajectories and their associations with PD risk and age at onset (AAO).

**Design:** Two ongoing online cohorts: Parkinson’s Progression Markers Initiative (PPMI)-Online (discovery; started in 2021) and Fox Insight (FI; validation; started in 2018).

**Setting:** Participants reported sleep duration across life stages, with PD-related follow-ups every three months.

**Participants:** A convenience sample of 5,660 individuals with PD and 10,245 without PD from PPMI-Online, and 1,929 participants with PD from FI.

**Exposures:** Self-reported sleep duration from ages 18 to 80+ in PPMI-Online and 12 to 66+ in FI. Latent class growth analysis (LCGA) identified sleep trajectories.

**Main Outcomes and Measures:** PD risk and AAO were assessed using logistic and linear regression, adjusting for demographics, lifestyle, comorbidities.

**Results:** The combined sample included 17,834 participants [age at sleep report 67.2±7.83 years; 9,735 (54.6%) female]. LCGA identified nine sleep trajectories in PPMI-Online, stable in early adulthood but diverging in midlife (stable, increasing, decreasing). Midlife sleep reductions (6-7 to ≤5-6 hours/day: OR = 1.90, 95% CI 1.61-2.24, *P* < .001; 7-8 to ≤6-7 hours/day: OR = 1.64, 95% CI 1.40-1.91, *P* < .001) and consistent short sleep (<=6 hours/day throughout adulthood: OR = 1.41, 95% CI 1.19-1.67, P < .001) demonstrated increased PD risk. Short sleep in early adulthood or midlife also had earlier AAO. The strongest effects were seen in those with ≤6 hours/day throughout adulthood (PPMI-Online: β = −2.45 years, 95% CI −3.33 to −1.56, P <.001) and those with a continuous decrease since adolescence (FI: β = −4.23 years, 95% CI −5.52 to −2.93, P < .001). These effects were independent of rapid eye movement sleep behavior disorders.

**Conclusions and Relevance:** Self-reported short sleep in early adulthood and midlife sleep reductions are associated with increased PD risk and earlier AAO. Self-perceived midlife sleep reduction may be a marker for future PD. Persons with chronic short sleep may be candidates for preventive intervention.

**Key Points:** **Question:** What is the relationship between life course sleep duration trajectories and the risk and age at onset of Parkinson’s disease (PD)?

**Findings:** In this cohort study including 7,589 participants with PD and 10,245 without PD, short sleep duration in early adulthood or decreasing sleep duration after age 50 were associated with an increased risk and earlier age at onset of PD.

**Meaning:** Short sleep duration over the life course, even in early adulthood (< 40 years), may contribute to risk of late-life PD; declining sleep duration after age 50 is a potential prodromal sign of PD.

## Introduction

Sleep disturbances are common nonmotor symptoms of Parkinson’s disease (PD), affecting 60% to 90% of patients.^1,2^ Evidence indicates a bidirectional relationship between PD and altered sleep, even before symptomatic onset.^3,4^ Sleep deprivation exacerbates synucleinopathy in rodent models,^5^ and poor sleep in human patients is associated with faster motor progression,^6^ indicating the role of sleep disturbances in PD pathogenesis. Furthermore, PD pathology disrupts the sleep/wake neural circuitry before significant dopaminergic neuron loss.^3^ This disruption is exemplified by rapid eye movement sleep behavioral disorder (RBD), a well-established prodromal marker of PD.^7^

Population-based studies report higher rates of insomnia or hypersomnia,^8–10^ and variations in sleep duration^11,12^ preceding PD diagnosis. However, these studies typically focus on isolated time points, primarily the decade before diagnosis, overlooking potential life course changes in sleep duration.^13^ Sleep trajectories have been associated with adverse neurological outcomes, such as dementia,^14–16^ but no study has examined the association between life course sleep duration and PD. Given that prodromal PD can span decades^17^ and early-life sleep patterns may contribute to neurodegeneration later in life,^18^ it is crucial to investigate early-life sleep trajectories and their connection to PD onset. Such research is critical for disentangling the role of sleep disturbances as prodromal or risk factors for PD, offering opportunities for early detection and intervention.

In this study, we identified self-reported sleep duration trajectories from adolesence to late adulthood using data from the Parkinson’s Progression Marker Initiative Online (PPMI-Online) and Fox Insight (FI). We assessed their relationship with PD risk and age at onset (AAO) in PPMI-Online, validating AAO findings in FI. We hypothesized that midlife sleep changes could serve as early markers of PD and that sleep durations below recommendations (<7 hours/night^19,20^) in early adulthood or midlife would be associated with higher risk and earlier AAO of PD. Sensitivity analyses excluded participants with self-reported RBD, anxiety or depression, dementia, and examined sex as a potential effect modifier.

## Methods

### PPMI-Online and FI Study Design

Both studies are ongoing longitudinal observational cohorts involving participants with and without PD. Convenience sampling was used to recruit participants aged 18 and older with minimal eligibility criteria. Recruitment was conducted through online channels, such as social media and email outreach, and on-site methods, including clinician referral and PD-related research events. Recruitment began in 2017 for FI^21,22^ and 2021 for PPMI-Online.^23^ Participants report PD status and symptoms quarterly via an online platform and complete occasional questionnaires, including those on sleep duration. The sleep questionnaire was administered during the 3rd and 15th months for PPMI-Online and between October 2017 and March 2019 for the FI study. For participants with multiple sleep reports, the most recent data were used for analysis. Study protocols were approved by institutional review boards (FI: New England IRB# 120160179; PPMI-Online: WCG IRB# 20211908), and all participants provided online informed consent. This study followed the Strengthening the Reporting of Observational Studies in Epidemiology (STROBE) reporting guideline.

### Study Participants

A flowchart of study participants is presented in Figure 1. The final sample in the discovery cohort included 11,728 participants from PPMI-Online (5,660 with PD and 10,245 without PD). Among the latter, 6,922 had complete data and were classified as “probable prodromal PD (pPD)” (n = 1,104) or “no probable pPD” (n = 5,818) based on the Movement Disorders Society criteria^24^ (80% probability cutoff^25^; **eMethods 1**). The validation FI sample included 1,929 participants with PD; the 338 participants without PD were excluded due to limited sample size.

**Figure 1.**
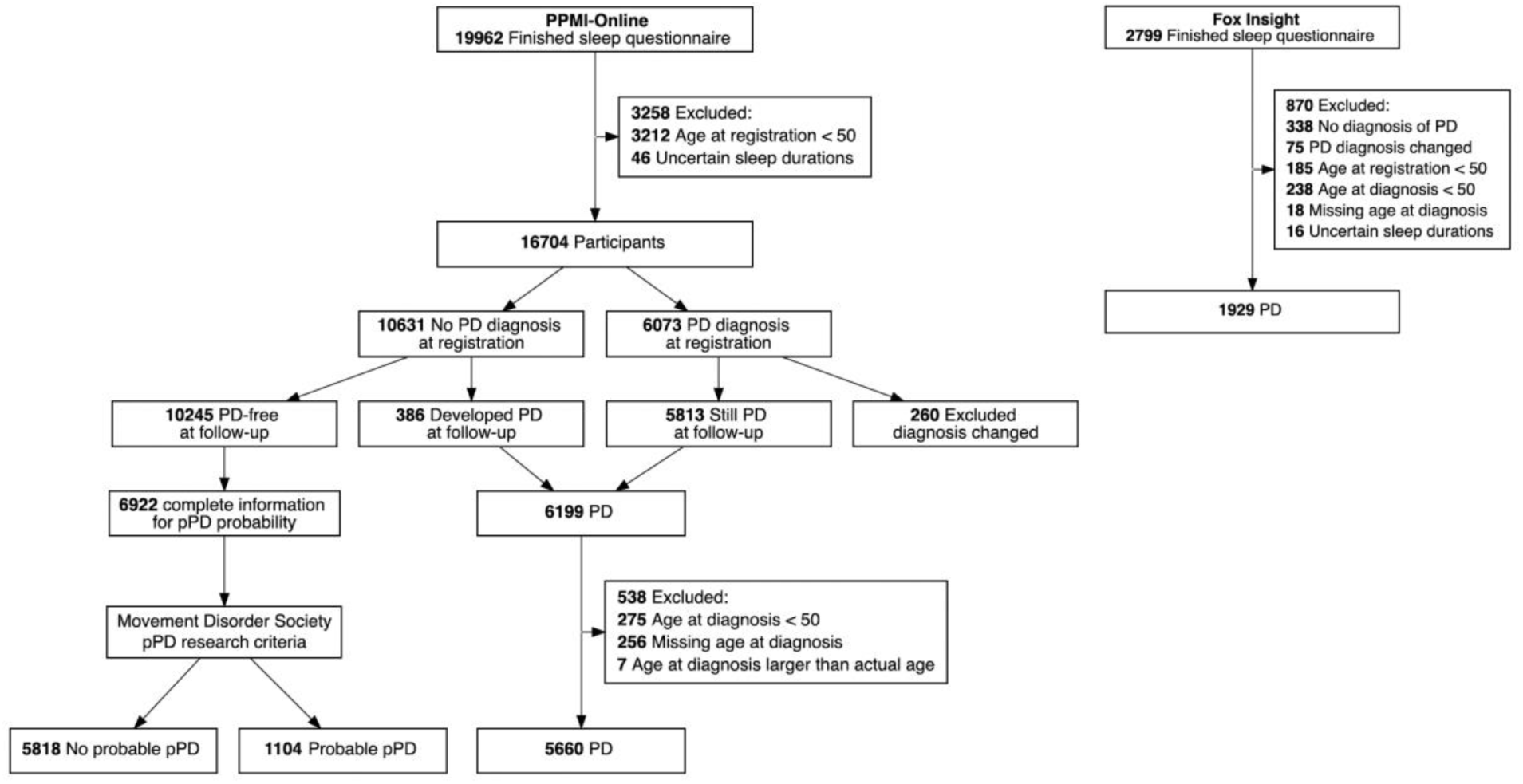
Flow Diagram of Study Participants. Abbreviations: PD = Parkinson’s disease, pPD = prodromal Parkinson’s disease

For the current analysis, we applied identical exclusion criteria to both cohorts, including age at registration < 50 years, young-onset PD (AAO < 50 years^26^), diagnosis changed during follow-up, missing age at diagnosis, reported age at diagnosis older than the participant’s current age, and all sleep duration responses marked as “prefer not to answer” or “don’t know”.

### Self-Perceived Sleep Duration Across the Lifespan

Participants reported their typical sleep duration during current and prior age periods. Response options were categorical and included: less than 5 hours, 5-6 hours, 6-7 hours, 7-8 hours, more than 8 hours, don’t know, and prefer not to answer. Responses of “don’t know” or “prefer not to answer” were treated as missing values.The PPMI-Online study captured data spanning adulthood, with more granular age categories during late adulthood (18-29, 30-39, 40-49, 50-59, 60-64, 65-69, 70-74, 75-79, and 80 or older). The FI study focused on earlier life stages (12-17, 18-25, 26-35, 36-45, 46-55, 56-65, and 66 or older), capturing transitions from adolescence to adulthood.

### Outcome Measures

PD status was self-reported at registration and follow-up, with previous studies confirming the reliability of self-reported PD in an online setting.^27^ The self-reported age at first PD diagnosis was used as a proxy for PD AAO, as these measures were highly correlated.^28^

### Statistical Analysis

We used latent class growth analysis (LCGA) to classify individuals into distinct sleep duration trajectory groups separately for PPMI-Online and FI, incorporating both linear and quadratic time components. LCGA identifies latent classes with similar trajectory patterns over time, assuming homogeneity within each class.^29^ In the initial phase, we identified trajectory groups in participants with complete sleep duration data from ages 18-29 to 75-79 for PPMI-Online (n = 2,643) and ages 12-17 to 56-65 for FI (n = 1,668), with the midpoint of each age range approximating age. We excluded the ‘80 or older’ and ‘66 or older’ groups due to limited sample sizes and broad age ranges. Participants lacking sleep data for all required periods were excluded to avoid overrepresentation of younger groups and sparse data for older groups. Those who selected “prefer not to answer” or “not sure” for any age period were also excluded to avoid potential bias from recall errors. In the next phase, all participants with at least one valid data point were assigned to the trajectory class with the highest membership probability. To assess trajectory consistency and robustness, we visually compared trajectory groups from (1) participants with complete data across age periods (main analysis) versus all participants, (2) those with PD versus without PD, and (3) those with RBD symptoms/diagnoses versus those without RBD. We tested models with 1 to 9 classes, selecting the optimal model based on lower Bayesian Information Criteria (BIC) and interpretability.^30^ BIC decreased as the number of classes increased from 1 to 9 for both PPMI-Online and FI. We selected the 9-class models for PPMI-Online and the 7-class model for FI, as additional classes showed similar patterns with no meaningful differences and limited sample size. Further LCGA details are provided in **eMethods 2**.

We analyzed associations between sleep duration trajectories and PD risk using logistic regression, and PD AAO using linear regression. Given possible non-monotonic changes during the prodromal phase,^31,32^ we performed seperate binomial logistic regressions for “PD” (with “No PD” as the refrence) and “probable pPD” (with “no probable pPD” as the reference). We built three models: Model 1 (unadjusted), Model 2 (adjusted for age at sleep report [for risk analyses only], sex, race, family history of PD, education, and income), and Model 3 (further adjusted for history of anxiety, depression, brain injury, diabetes, hypertension; RBD Single-Question Screen^33^ at the time of sleep report; lifetime caffeine intake, smoking status, and lifetime physical inactivity). We did not include age at sleep report as a covariate for AAO as AAO typically precedes the sleep report and is minimally influenced by the age at sleep report. Covariates, selected based on their known or suspected associations with sleep and PD,^34^ were self-reported, with the closest report to sleep report used for repeated measures. All models met assumptions, with no variance inflation factor exceeding 5, indicating minimal multicollinearity.

Sensitivity analyses included: (1) restricting to participants without self-reported RBD (n = 8395), given its association with shorter sleep,^35^ (2) restricting to participants without history of anxiety or depression (n = 8566), given the divergent effects of these conditions and their treatments on sleep, (3) excluding participants with possible dementia (Penn Parkinson’s Daily Activities Questionnaire-15 < 43^36^) to enhance recall accuracy (n = 14405), and (4) exploring sex as a possible effect modifier. Furthermore, as baseline sleep during early adulthood and trends during midlife are critical components of the sleep duration trajectories, we also investigated whether these two factors are independently associated with PD risk and AAO.

We performed data analysis from January to December 2024 using R (version 4.3.2). Major packages included “lcmm”^37^ (version 2.1.0), “stats” (version 4.3.2), “ggplot2” (version 3.4.4). All tests were 2-sided, with statistical significance set at *P* < .05.

## Results

### Study Participants

Demographic and clinical characteristics of participants is summarized in Table 1: 15,905 from PPMI-Online [mean age at sleep report: 67.2±7.94 years; 8,795 (55.3%) female; mean follow-up: 22.7±8.71 months] and 1,929 from FI [mean age at sleep report: 67.5±6.87 years; 940 (48.7%) female; mean follow-up: 68.1±10.1 months]. PD mean AAO was 65.3±7.57 in PPMI-Online and 62.9±6.96 in FI. In PPMI-Online, participants without PD were younger, more likely female, and had higher education, income, and positive PD family history compared to those with PD. Across subgroups (“no probable pPD,” “probable pPD,” and “PD”; **eTable 1**), motor and non-motor symptoms showed a clear progression, supporting the validity of the pPD criteria.

**Table 1.**
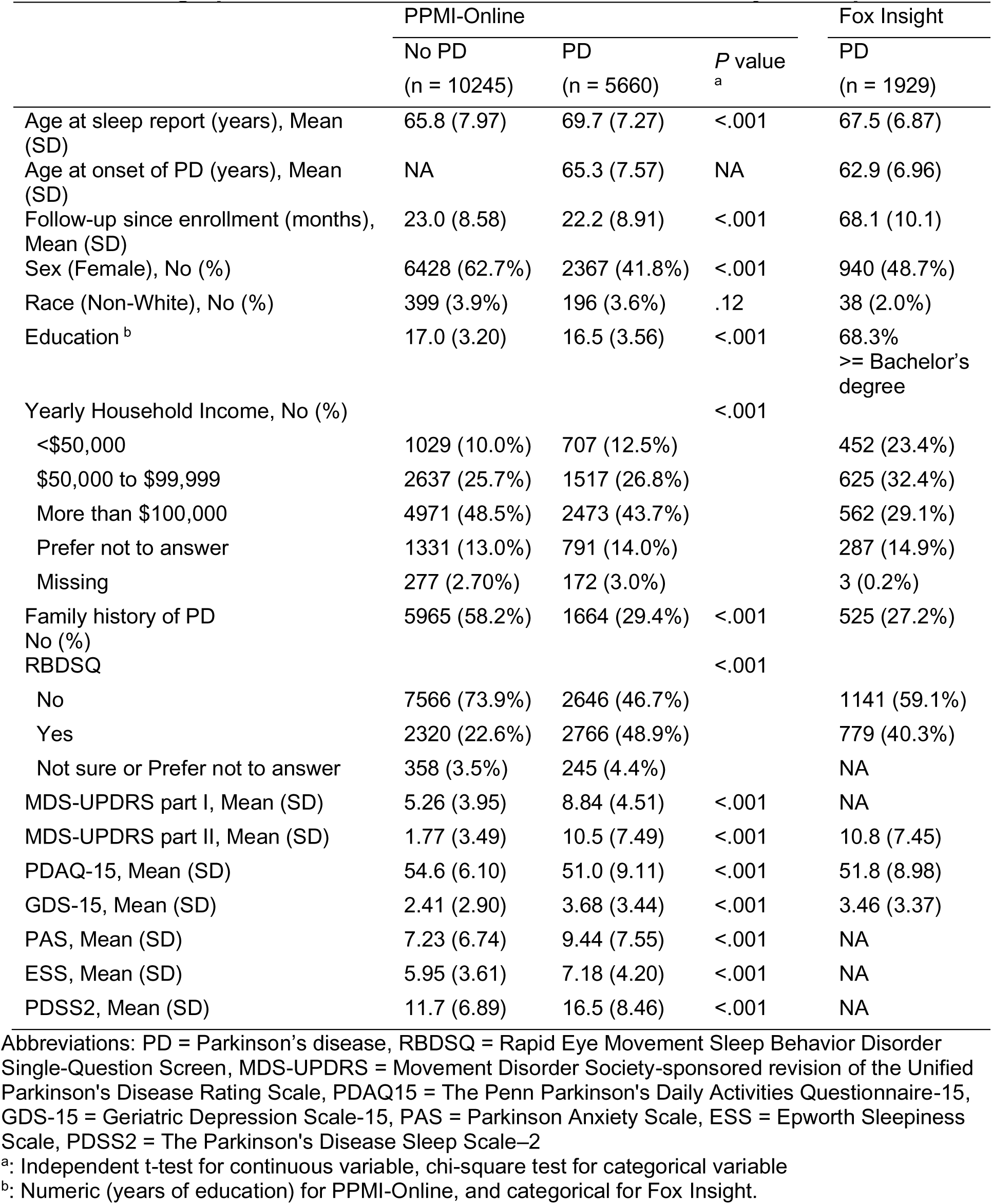
Demographic and Clinical Characteristics of the Study Participants.

### Sleep Duration Trajectories (PPMI-Online)

PPMI-Online sleep duration trajectories (**Figure 2A**) remained relatively stable from the 20s to the 40s, with distinct trends emerging after the 50s (depicted in separate panels). Groups were labeled according to both baseline sleep in early adulthood (with categories: >8 hours, 7-8 hours, 6-7 hours, <=6 hours) and changes after age 50 (categorized as: stable, decrease, or increase). These trajectory shapes were largely consistent in sensitivity analyses. The “<=6 hrs stable” group had a lower age at sleep report, smaller percentage of White individuals, fewer years of education, and lower household income. Groups with changes after 50s (“7-8 hrs increase”, “6-7 hrs decrease”, “7-8 hrs decrease”) had higher age at sleep report and lower income. The “>8 hrs stable” group included more females (**eTable 2**).

**Figure 2.**
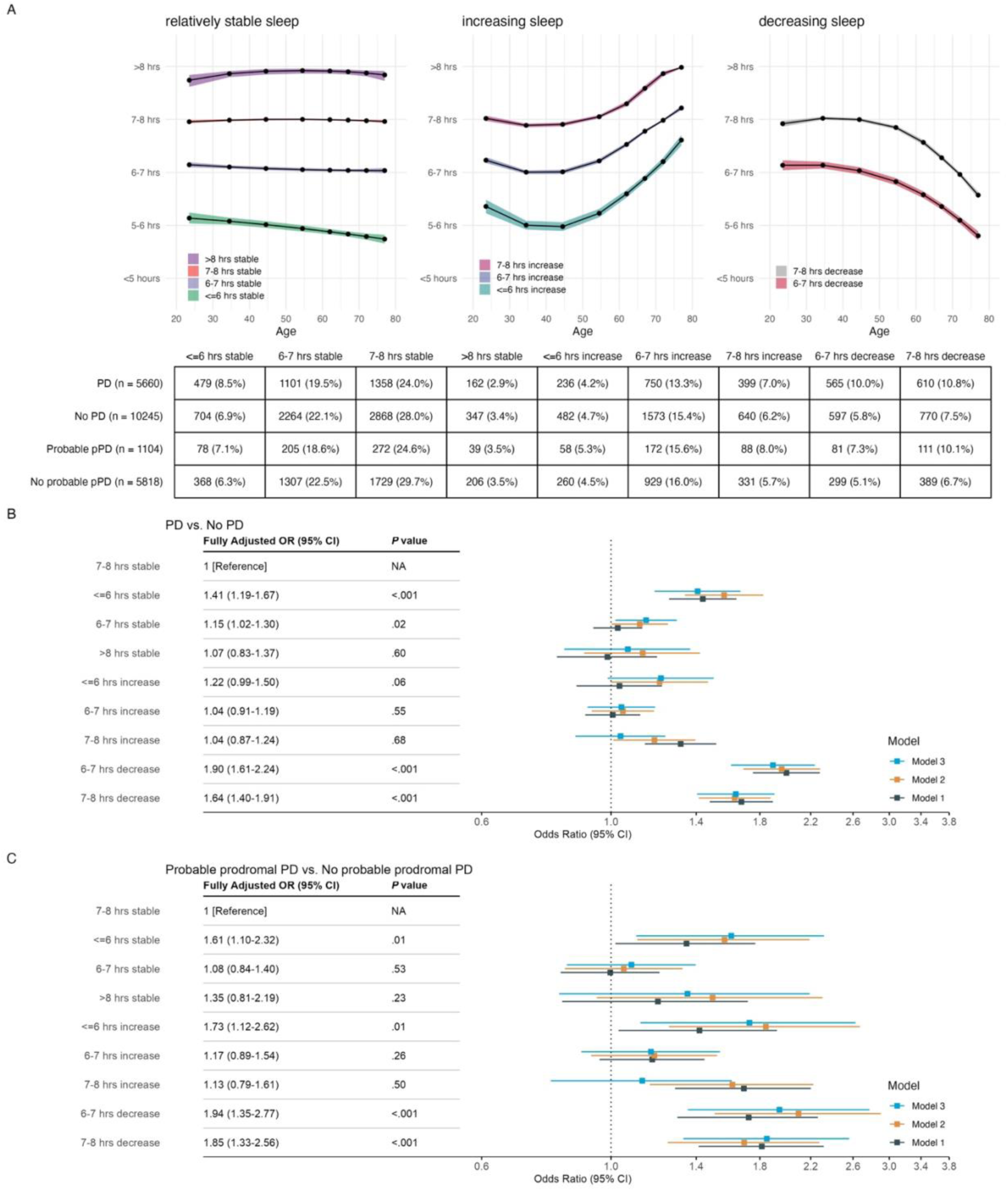
Sleep Duration Trajectories Identified from PPMI-Online and Associations with Parkinson’s Disease Risk. Abbreviations: PD = Parkinson’s disease, pPD = prodromal Parkinson’s disease Panel A: Sleep duration trajectory groups identified through latent class growth analysis, shown in separate panels based on trends emerging after age 50. Dots and shaded areas represent mean trajectories with 95% confidence intervals. The accompanying table shows the count and proportion of participants for each trajectory pattern. Panel B: Logistic regression results for the association between sleep duration trajectory patterns and risk of PD, using participants without a PD diagnosis as the reference group. Panel C: Logistic regression of sleep duration trajectory patterns and risk of “Probable pPD”, with the “No probable pPD” group as the reference. Model 1 was unadjusted. Model 2 adjusted for age at sleep report, sex, race, family history of PD, education, and income. Model 3 further adjusted for history of anxiety, depression, brain injury, diabetes, hypertension; Rapid Eye Movement Sleep Behavior Disorder Single-Question Screen at the time of sleep report; lifetime caffeine intake, smoking status, and lifetime physical inactivity.

### Sleep Duration Trajectories and PD Risk (PPMI-Online)

Compared to the “7-8 hours stable” group, those with consistently shorter sleep (“<=6 hours stable”: OR = 1.41, 95% CI 1.19-1.67, *P* < .001, all ORs from Model 3) or decreasing sleep after age 50 (“6-7 hours decrease”: OR = 1.90, 95% CI 1.61-2.24, *P* < .001, “7-8 hours decrease”: OR = 1.64, 95% CI 1.40-1.91, *P* < .001) showed increased PD risk. The associations remained consistent across all adjustments (**Figure 2B**). The “6-7 hrs stable” group also showed a modestly higher risk of PD after adjustment (OR = 1.15, 95% CI 1.02-1.30, *P* = .02).

For probable pPD, consistently shorter sleep (“<=6 hours stable”: OR = 1.61, 95% CI 1.10-2.32, *P* = .01) and decreasing sleep after 50 (“6-7 hours decrease”: OR = 1.94, 95% CI 1.35-2.77, *P* < .001; “7-8 hours decrease”: OR = 1.85, 95% CI 1.33-2.56, *P* < .001) were similarly associated with higher risks, consistent with the findings for PD risk. In addition, the “<=6 hrs increase” group also exhibited elevated pPD risk (OR = 1.73, 95% CI 1.12-2.62, *P* = .01).

### Sleep Duration Trajectories and PD AAO (PPMI-Online)

Compared to the “7-8 hours stable” group, short sleep in early adulthood or decreasing sleep in midlife was associated with earlier PD AAO (**Figure 3**). After full adjustment, the largest effect size was observed in the “<=6 hrs stable” group (β = −2.45 years, 95% CI −3.33 to −1.56, *P* = <.001) and “6-7 hrs decrease” group (β = −2.37 years, 95% CI −3.19 to −1.56, *P* = <.001), followed by the “<=6 hrs increase” (β = −1.73 years, 95% CI −2.85 to −0.62, *P* = .002) and “6-7 hrs stable” (β = −0.91 years, 95% CI −1.57 to −0.26, *P* = .006).

**Figure 3.**
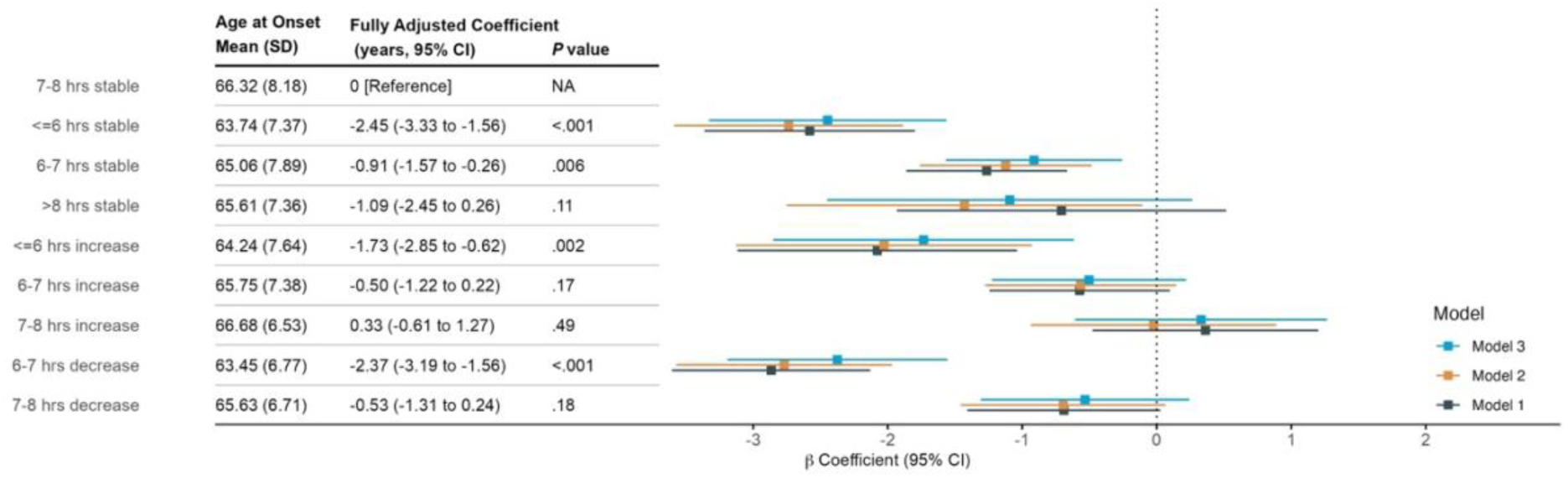
Association of Life Course Sleep Duration with Age at Onset of Parkinson’s Disease in PPMI-Online. Model 1 was unadjusted. Model 2 adjusted for sex, race, family history of PD, education, and income. Model 3 further adjusted for history of anxiety, depression, brain injury, diabetes, hypertension; Rapid Eye Movement Sleep Behavior Disorder Single-Question Screen at the time of sleep report; lifetime caffeine intake, smoking status, and lifetime physical inactivity.

### Sleep Duration Trajectories and Associations with PD AAO (FI)

Most FI trajectories (**Figure 4A**) exhibited moderate sleep decline from adolescence to adulthood. Among them, four groups (labeled “>8 hours”, “7-8 hours”, “6-7 hours”, “<=6 hours”) maintained relatively stable during midlife. Two groups showed continuous declines: “decrease-1,” with decreases earlier in life, and “decrease-2,” with declines more evident during midlife. The “increase” group exhibited a sleep increase after the 40s, with early adulthood sleep duration between 6-7 hours, resembling the “6-7 hours increase” group in PPMI-Online. Groups with short sleep (“<=6 hrs”, “6-7 hrs”) and decreasing sleep (“decrease-1”, “decrease-2”) had lower education levels. The decrease groups also had a younger age at sleep report (**eTable 3**).

**Figure 4.**
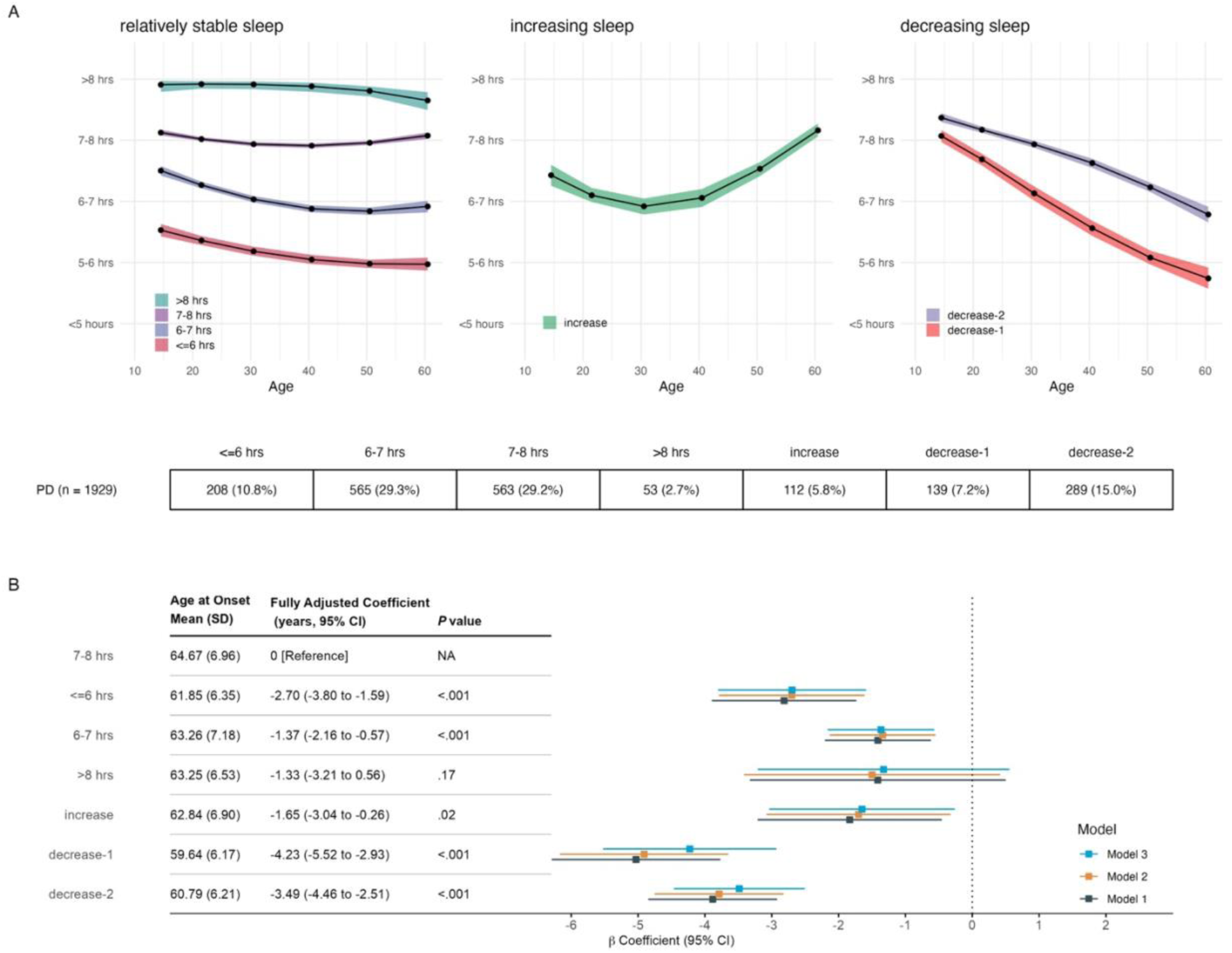
Sleep Duration Trajectories Identified from Fox Insight and Associations with Parkinson’s Disease Age at Onset. Panel A: Dots and shaded areas represent mean trajectories for each class with 95% confidence intervals. The table displays the count and proportion of each trajectory pattern. Panel B: Sleep duration trajectory patterns and age at onset of PD. Model 1 was unadjusted. Model 2 adjusted for sex, race, family history of PD, education, and income. Model 3 further adjusted for history of anxiety, depression, brain injury, diabetes, hypertension; Rapid Eye Movement Sleep Behavior Disorder Single-Question Screen at the time of sleep report; lifetime caffeine intake, smoking status, and lifetime physical inactivity.

Compared to the “7-8 hours” group, groups with less sleep exhibited earlier AAO (**Figure 4B**). The effect size was most pronounced in the decrease groups (“decrease-1”: β = −4.23 years, 95% CI −5.52 to −2.93, *P* < .001; “decrease-2” (β = −3.49 years, 95% CI −4.46 to −2.51, *P* < .001), followed by “<=6 hours” (β = −2.70 years, 95% CI −3.80 to −1.59, *P* < .001), “increase” (β = −1.65 years, 95% CI −3.04 to −0.26, *P* = .02), and “6-7 hours” (β = −1.37 years, 95% CI −2.16 to −0.57, *P* < .001).

### Sensitivity Analyses

The associations remained consistent across sexes and were robust among participants without RBD symptoms or diagnoses throughout follow-up, those without a history of anxiety or depression, and after excluding individuals with possible dementia (**eFigure 1-6**). Additionally, shorter baseline sleep duration and decreasing sleep during midlife were independently linked to an increased PD risk and earlierer AAO (**eFigure 7**).

## Discussion

In two large PD-centered online cohorts, we identified distinct self-reported life course sleep duration trajectories and examined their associations with PD risk and AAO. PPMI-Online results highlighted midlife, particularly around age 50, as a critical turning point in sleep duration. Declining sleep during midlife was associated with up to a 90% higher risk of PD and pPD. Chronic short sleep from early adulthood was associated with a 1.4-fold increased PD risk. Additionally, both PPMI-Online and FI cohorts indicated that shorter-than-recommended sleep in early adulthood or midlife was associated with an earlier AAO, with such patterns associated with earlier AAO by up to 2.5 to 4.2 years.

Due to the different life stage focuses, PPMI-Online and FI trajectories exhibited complementary patterns. Around 50% of PPMI-Online participants were in trajectory groups showing changes after age 50, and 60% of FI participants belong to groups showing declines from adolescence to the 30s. These findings align with previously identified turning points in sleep duration at ages 33 and 53.^13^ However, stable groups might be overestimated due to categorical presentation, potentially missing subtle changes.

Midlife (ages 40s to 60s) is increasingly recognized as a critical period marked by accelerated declines in gene expressions, brain connectivity, and cognitive function.^38,39^ We observed that decreasing sleep after age 50 was associated with increased PD and pPD risk, suggesting that declining sleep is evident in prodromal PD and continues into clinical PD. This association remained robust even among participants without self-reported RBD, anxiety, or depression, indicating that decreasing sleep may be an early manifestation of PD itself, rather than a consequence of common prediagnostic comorbidities observed in a subset of patients.^40^ Conversely, the “≤6 hrs increase” group showed an increased risk of pPD but not PD, suggesting that increasing sleep during midlife, particularly after chronic short sleep, may indicate prodromal PD. However, our study lacked an ultra-long sleep group (>8 hours/day), limiting comparisons to previous reports linking >=9 hours/day with higher PD risk.^41,42^

Given the long prodromal window, it is challenging to determine the direction of the relationship between sleep disturbances and PD. Our study indicates that short sleep in early adulthood or midlife is associated with increased risk and earlier AAO of PD, potentially preceding the onset of PD by up to five decades. These findings suggest that chronic short sleep might be a risk factor, with prolonged short sleep likely preceding neurodegeneration. Our results also underscore the public health relevance of addressing insufficient sleep, particularly in minoritized populations,^43^ and its adverse effects on aging-related outcomes.^44^ As preventive trials for PD are being planned,^45^ promoting healthy sleep hygiene may represent a scalable strategy for reducing risk or delaying onset.

Our findings align with mechanistic evidence from animal and human studies. Sleep disturbances may arise from early PD pathology affecting the locus coeruleus,^46^ hypothalamus,^47^ and dopaminergic systems.^48^ Conversely, sleep deprivation exacerbates synucleinopathy,^5^ possibly through mitochondrial and oxidative stress,^49^ neuroinflammation,^50^ impaired glymphatic clearance,^51^ and synaptic homeostasis.^52^ Notably, the locus coeruleus is particularly vulnerable to insufficient sleep and may contribute to PD pathogenesis.^53,54^ Furthermore, chronic sleep loss in young adult rodents has long-lasting neurological effects^55^ and is associated with neurodegeneration later in life,^18^ supporting our findings that short sleep in early adulthood might contribute to PD in later years.

This study leveraged two online PD-centered cohorts, providing a cost-efficient framework with a large participant pool and longitudinal sleep duration data spanning adolescence to age 80. However, several limitations should be considered. First, the retrospective self-reported sleep duration may introduce recall bias, although evidence suggests such information can be accurate.^56,57^ Self-reported sleep duration modestly correlates with objectively measured sleep.^58^ In this study, participants could select “don’t know” for uncertain responses, and results remained robust after excluding possible dementia. Additionally, the sleep trajectory turning points aligned with a previous large-scale study,^13^ enhancing the credibility of self-reported sleep. Even if discrepancies exist, self-perceived decline in sleep after midlife may still serve as a valuable indicator of PD risk. Second, pPD was determined by self-reported markers, without clinical evaluation or synuclein-based biomarkers. Furthermore, the study population was predominantly White, with high socioeconomic status and education, and participants without PD were enriched for PD risk factors. Further studies are needed to assess generalizability. Finally, while the findings suggest short sleep as a potential PD risk factor, the observational nature of this study limits causal inferences.

## Conclusion

In this large-scale study involving individuals with and without PD, we found that the 50s represent a significant turning point in sleep duration. Monitoring sleep duration changes, particularly after age 50, is crucial, as declining sleep duration at this stage may signal a higher risk and earlier onset of PD. Additionally, chronic short sleep was associated with an increased risk and earlier AAO of PD, suggesting short sleep as a potential risk factor and intervention targets.

## Data Availability

All data produced are available online at www.ppmi-info.org and https://foxinsightinfo.michaeljfox.org/insight/explore/insight.jsp

https://foxden.michaeljfox.org/insight/explore/insight.jsp

## Acknowledgement

We would like to thank the Parkinson’s community for participating in the Fox Insight and PPMI-Online study to make this research possible.

The Fox Insight Study is funded by The Michael J. Fox Foundation for Parkinson’s Research.

PPMI – a public-private partnership – is funded by the Michael J. Fox Foundation for Parkinson’s Research and funding partners, including 4D Pharma, Abbvie, AcureX, Allergan, Amathus Therapeutics, Aligning Science Across Parkinson’s, AskBio, Avid Radiopharmaceuticals, BIAL, Biogen, Biohaven, BioLegend, BlueRock Therapeutics, Bristol-Myers Squibb, Calico Labs, Celgene, Cerevel Therapeutics, Coave Therapeutics, DaCapo Brainscience, Denali, Edmond J. Safra Foundation, Eli Lilly, Gain Therapeutics, GE HealthCare, Genentech, GSK, Golub Capital, Handl Therapeutics, Insitro, Janssen Neuroscience, Lundbeck, Merck, Meso Scale Discovery, Mission Therapeutics, Neurocrine Biosciences, Pfizer, Piramal, Prevail Therapeutics, Roche, Sanofi, Servier, Sun Pharma Advanced Research Company, Takeda, Teva, UCB, Vanqua Bio, Verily, Voyager Therapeutics, the Weston Family Foundation and Yumanity Therapeutics.

## Data Sharing Statement

Final data used in this article were obtained from the PPMI study (tier1 data downloaded on 03 July 2024, https://www.ppmi-info.org/access-data-specimens/download-data, RRID:SCR_006431) and the FI study (version: December 2024 archived monthly data, https://foxinsight-info.michaeljfox.org/insight/explore/insight.jsp). For up-todate information on the PPMI study, visit www.ppmi-info.org. For up-to-date information on the FI study, visit https://foxinsightinfo.michaeljfox.org/insight/explore/insight.jsp.

## References

1. Lajoie AC, Lafontaine AL, Kaminska M. The Spectrum of Sleep Disorders in Parkinson Disease: A Review. Chest. 2021;159(2):818–827. doi:10.1016/j.chest.2020.09.099

2. Iranzo A, Cock VCD, Fantini ML, Pérez-Carbonell L, Trotti LM. Sleep and sleep disorders in people with Parkinson’s disease. The Lancet Neurology. 2024;0(0). doi:10.1016/S1474-4422(24)00170-4

3. Al-Qassabi A, Fereshtehnejad SM, Postuma RB. Sleep Disturbances in the Prodromal Stage of Parkinson Disease. Curr Treat Options Neurol. 2017;19(6):22. doi:10.1007/s11940-017-0458-1

4. Leng Y, Blackwell T, Cawthon PM, Ancoli-Israel S, Stone KL, Yaffe K. Association of Circadian Abnormalities in Older Adults With an Increased Risk of Developing Parkinson Disease. JAMA Neurology. 2020;77(10):1270–1278. doi:10.1001/jamaneurol.2020.1623

5. Morawska MM, Moreira CG, Ginde VR, et al. Slow-wave sleep affects synucleinopathy and regulates proteostatic processes in mouse models of Parkinson’s disease. Science Translational Medicine. 2021;13(623):eabe7099. doi:10.1126/scitranslmed.abe7099

6. Schreiner SJ, Imbach LL, Werth E, et al. Slow-wave sleep and motor progression in Parkinson disease. Annals of Neurology. 2019;85(5):765–770. doi:10.1002/ana.25459

7. Postuma RB, Iranzo A, Hu M, et al. Risk and predictors of dementia and parkinsonism in idiopathic REM sleep behaviour disorder: a multicentre study. Brain. 2019;142(3):744–759. doi:10.1093/brain/awz030

8. Schrag A, Horsfall L, Walters K, Noyce A, Petersen I. Prediagnostic presentations of Parkinson’s disease in primary care: a case-control study. Lancet Neurol. 2015;14(1):57–64. doi:10.1016/S1474-4422(14)70287-X

9. Simonet C, Bestwick J, Jitlal M, et al. Assessment of Risk Factors and Early Presentations of Parkinson Disease in Primary Care in a Diverse UK Population. JAMA Neurol. 2022;79(4):359. doi:10.1001/jamaneurol.2022.0003

10. Schrag A, Bohlken J, Dammertz L, et al. Widening the Spectrum of Risk Factors, Comorbidities, and Prodromal Features of Parkinson Disease. JAMA Neurol. 2023;80(2):161–171. doi:10.1001/jamaneurol.2022.3902

11. Otaiku AI. Association of sleep abnormalities in older adults with risk of developing Parkinson’s disease. Sleep. 2022;45(11):zsac206. doi:10.1093/sleep/zsac206

12. Lysen TS, Darweesh SKL, Ikram MK, Luik AI, Ikram MA. Sleep and risk of parkinsonism and Parkinson’s disease: a population-based study. Brain. 2019;142(7):2013–2022. doi:10.1093/brain/awz113

13. Coutrot A, Lazar AS, Richards M, et al. Reported sleep duration reveals segmentation of the adult life-course into three phases. Nat Commun. 2022;13(1):7697. doi:10.1038/s41467-022-34624-8

14. Cavaillès C, Carrière I, Wagner M, et al. Trajectories of sleep duration and timing before dementia: a 14-year follow-up study. Age Ageing. 2022;51(8):afac186. doi:10.1093/ageing/afac186

15. Lu Y, Sugawara Y, Zhang S, Tomata Y, Tsuji I. Changes in sleep duration and the risk of incident dementia in the elderly Japanese: the Ohsaki Cohort 2006 Study. Sleep. 2018;41(10):zsy143. doi:10.1093/sleep/zsy143

16. Sabia S, Fayosse A, Dumurgier J, et al. Association of sleep duration in middle and old age with incidence of dementia. Nat Commun. 2021;12(1):2289. doi:10.1038/s41467-021-22354-2

17. Berg D, Borghammer P, Fereshtehnejad SM, et al. Prodromal Parkinson disease subtypes — key to understanding heterogeneity. Nat Rev Neurol. 2021;17(6):349–361. doi:10.1038/s41582-021-00486-9

18. Owen JE, Zhu Y, Fenik P, et al. Late-in-life neurodegeneration after chronic sleep loss in young adult mice. Sleep. 2021;44(8):zsab057. doi:10.1093/sleep/zsab057

19. Watson NF, Badr MS, Belenky G, et al. Recommended Amount of Sleep for a Healthy Adult: A Joint Consensus Statement of the American Academy of Sleep Medicine and Sleep Research Society. SLEEP. Published online June 1, 2015. doi:10.5665/sleep.4716

20. Hirshkowitz M, Whiton K, Albert SM, et al. National Sleep Foundation’s updated sleep duration recommendations: final report. Sleep Health. 2015;1(4):233–243. doi:10.1016/j.sleh.2015.10.004

21. Smolensky L, Amondikar N, Crawford K, et al. Fox Insight collects online, longitudinal patient-reported outcomes and genetic data on Parkinson’s disease. Sci Data. 2020;7(1):67. doi:10.1038/s41597-020-0401-2

22. Gottesman J, Karim Y, Forbes J, et al. Fox Insight at 5 years - a cohort of 54,000 participants contributing longitudinal patient-reported outcome, genetic, and microbiome data relating to Parkinson’s disease. Sci Data. 2024;11(1):615. doi:10.1038/s41597-024-03407-9

23. M Tanner C. Parkinson’s Progression Markers Initiative Online Study (PPMI Online) v1. Published online March 8, 2024. doi:10.17504/protocols.io.q26g718y9gwz/v1

24. Heinzel S, Berg D, Gasser T, et al. Update of the MDS research criteria for prodromal Parkinson’s disease. Movement Disorders. 2019;34(10):1464–1470. doi:10.1002/mds.27802

25. Berg D, Postuma RB, Adler CH, et al. MDS research criteria for prodromal Parkinson’s disease. Movement Disorders. 2015;30(12):1600–1611. doi:10.1002/mds.26431

26. Mehanna R, Smilowska K, Fleisher J, et al. Age Cutoff for Early-Onset Parkinson’s Disease: Recommendations from the International Parkinson and Movement Disorder Society Task Force on Early Onset Parkinson’s Disease. Movement Disorders Clinical Practice. 2022;9(7):869–878. doi:10.1002/mdc3.13523

27. Myers TL, Tarolli CG, Adams JL, et al. Video-based Parkinson’s disease assessments in a nationwide cohort of Fox Insight participants. Clinical Parkinsonism & Related Disorders. 2021;4:100094. doi:10.1016/j.prdoa.2021.100094

28. Pihlstrøm L, Fan CC, Frei O, et al. Genetic Stratification of Age-Dependent Parkinson’s Disease Risk by Polygenic Hazard Score. Movement Disorders. 2022;37(1):62–69. doi:10.1002/mds.28808

29. Jung T, Wickrama K a. S. An Introduction to Latent Class Growth Analysis and Growth Mixture Modeling. Social and Personality Psychology Compass. 2008;2(1):302–317. doi:10.1111/j.1751-9004.2007.00054.x

30. Lennon H, Kelly S, Sperrin M, et al. Framework to construct and interpret latent class trajectory modelling. BMJ Open. 2018;8(7):e020683. doi:10.1136/bmjopen-2017-020683

31. Postuma RB, Gagnon JF, Pelletier A, Montplaisir JY. Insomnia and somnolence in idiopathic RBD: a prospective cohort study. npj Parkinson’s Disease. 2017;3(1):1–6. doi:10.1038/s41531-017-0011-7

32. Joza S, Hu MT, Jung KY, et al. Progression of clinical markers in prodromal Parkinson’s disease and dementia with Lewy bodies: a multicentre study. Brain. 2023;146(8):3258–3272. doi:10.1093/brain/awad072

33. Postuma RB, Arnulf I, Hogl B, et al. A single-question screen for rapid eye movement sleep behavior disorder: A multicenter validation study. Movement Disorders. 2012;27(7):913–916. doi:10.1002/mds.25037

34. Ben-Shlomo Y, Darweesh S, Llibre-Guerra J, Marras C, Luciano MS, Tanner C. The epidemiology of Parkinson’s disease. The Lancet. 2024;403(10423):283–292. doi:10.1016/S0140-6736(23)01419-8

35. Zhang Y, Ren R, Yang L, Sanford LD, Tang X. Polysomnographically measured sleep changes in idiopathic REM sleep behavior disorder: A systematic review and meta-analysis. Sleep Medicine Reviews. 2020;54:101362. doi:10.1016/j.smrv.2020.101362

36. Brennan L, Siderowf A, Rubright JD, et al. The Penn Parkinson’s Daily Activities Questionnaire-15: Psychometric properties of a brief assessment of cognitive instrumental activities of daily living in Parkinson’s disease. Parkinsonism & Related Disorders. 2016;25:21–26. doi:10.1016/j.parkreldis.2016.02.020

37. Proust-Lima C, Philipps V, Liquet B. Estimation of Extended Mixed Models Using Latent Classes and Latent Processes: The R Package lcmm. Journal of Statistical Software. 2017;78:1–56. doi:10.18637/jss.v078.i02

38. Dohm-Hansen S, English JA, Lavelle A, Fitzsimons CP, Lucassen PJ, Nolan YM. The “middle-aging” brain. Trends in Neurosciences. 2024;47(4):259–272. doi:10.1016/j.tins.2024.02.001

39. Yaffe K, Bahorik AL, Hoang TD, et al. Cardiovascular risk factors and accelerated cognitive decline in midlife: The CARDIA Study. Neurology. 2020;95(7). doi:10.1212/WNL.0000000000010078

40. Blesa J, Foffani G, Dehay B, Bezard E, Obeso JA. Motor and non-motor circuit disturbances in early Parkinson disease: which happens first? Nat Rev Neurosci. 2022;23(2):115–128. doi:10.1038/s41583-021-00542-9

41. Beydoun HA, Naughton MJ, Beydoun MA, et al. Association of sleep disturbance with Parkinson disease: evidence from the Women’s Health Initiative. Menopause. 2022;29(3):255–263. doi:10.1097/GME.0000000000001918

42. Chen Y, Gao Y, Sun X, et al. Association between Sleep Factors and Parkinson’s Disease: A Prospective Study Based on 409,923 UK Biobank Participants. Neuroepidemiology. 2023;57(5):293–303. doi:10.1159/000530982

43. Jean-Louis G, Grandner MA, Seixas AA. Social determinants and health disparities affecting sleep. The Lancet Neurology. 2022;21(10):864–865. doi:10.1016/S1474-4422(22)00347-7

44. Leng Y, Knutson K, Carnethon MR, Yaffe K. Association Between Sleep Quantity and Quality in Early Adulthood With Cognitive Function in Midlife. Neurology. 2024;102(2):e208056. doi:10.1212/WNL.0000000000208056

45. Berg D, Crotty GF, Keavney JL, Schwarzschild MA, Simuni T, Tanner C. Path to Parkinson Disease Prevention. Neurology. 2022;99(7_Supplement_1):76–83. doi:10.1212/WNL.0000000000200793

46. Braak E, Sandmann-Keil D, Rüb U, et al. α-Synuclein immunopositive Parkinson’s disease-related inclusion bodies in lower brain stem nuclei. Acta Neuropathol. 2001;101(3):195–201. doi:10.1007/s004010000247

47. De Pablo-Fernández E, Warner TT. Hypothalamic α-synuclein and its relation to autonomic symptoms and neuroendocrine abnormalities in Parkinson disease. In: Handbook of Clinical Neurology. Vol 182. Elsevier; 2021:223–233. doi:10.1016/B978-0-12-819973-2.00015-0

48. Holst SC, Bersagliere A, Bachmann V, Berger W, Achermann P, Landolt HP. Dopaminergic Role in Regulating Neurophysiological Markers of Sleep Homeostasis in Humans. J Neurosci. 2014;34(2):566–573. doi:10.1523/JNEUROSCI.4128-13.2014

49. Zamore Z, Veasey SC. Neural consequences of chronic sleep disruption. Trends in Neurosciences. 2022;45(9):678–691. doi:10.1016/j.tins.2022.05.007

50. Zhu B, Dong Y, Xu Z, et al. Sleep disturbance induces neuroinflammation and impairment of learning and memory. Neurobiology of Disease. 2012;48(3):348–355. doi:10.1016/j.nbd.2012.06.022

51. Xie L, Kang H, Xu Q, et al. Sleep Drives Metabolite Clearance from the Adult Brain. Science. 2013;342(6156):373–377. doi:10.1126/science.1241224

52. Zahed H, Zuzuarregui JRP, Gilron R, Denison T, Starr PA, Little S. The Neurophysiology of Sleep in Parkinson’s Disease. Movement Disorders. 2021;36(7):1526–1542. doi:10.1002/mds.28562

53. Butkovich LM, Houser MC, Chalermpalanupap T, et al. Transgenic Mice Expressing Human α-Synuclein in Noradrenergic Neurons Develop Locus Ceruleus Pathology and Nonmotor Features of Parkinson’s Disease. J Neurosci. 2020;40(39):7559–7576. doi:10.1523/JNEUROSCI.1468-19.2020

54. Ray Chaudhuri K, Leta V, Bannister K, Brooks DJ, Svenningsson P. The noradrenergic subtype of Parkinson disease: from animal models to clinical practice. Nat Rev Neurol. 2023;19(6):333–345. doi:10.1038/s41582-023-00802-5

55. Yuan RK, Lopez MR, Ramos-Alvarez MM, et al. Differential effect of sleep deprivation on place cell representations, sleep architecture, and memory in young and old mice. Cell Reports. 2021;35(11):109234. doi:10.1016/j.celrep.2021.109234

56. Berney LR, Blane DB. Collecting retrospective data: Accuracy of recall after 50 years judged against historical records. Social Science & Medicine. 1997;45(10):1519–1525. doi:10.1016/S0277-9536(97)00088-9

57. Havari E, Mazzonna F. Can We Trust Older People’s Statements on Their Childhood Circumstances? Evidence from SHARELIFE. Eur J Population. 2015;31(3):233–257. doi:10.1007/s10680-014-9332-y

58. Lauderdale DS, Knutson KL, Yan LL, Liu K, Rathouz PJ. Self-reported and measured sleep duration: how similar are they? Epidemiology. 2008;19(6):838–845. doi:10.1097/EDE.0b013e318187a7b0

